# Cardiovascular and Renal Outcomes among Patients with Type 2 Diabetes using SGLT2 Inhibitors added to Metformin: A Population-Based Cohort Study from the United Kingdom

**DOI:** 10.1101/2022.07.28.22278158

**Authors:** Antonio González-Pérez, David Vizcaya, María E Sáez, Marcus Lind, Luis A Garcia Rodriguez

## Abstract

**Introduction:** As large numbers of patients with type 2 diabetes receive treatment with a sodium-glucose co-transporter-2 inhibitor (SGLT2i), we investigated whether the cardio-renal preventative effects found in clinical trials are also seen in clinical practice where patient characteristics and adherence to treatment differs.

**Research design and methods:** Using UK primary care electronic health records, we followed two cohorts of patients with type 2 diabetes prescribed metformin: SGLT2is (N=12,978) and a matched comparator of patients not using a SGLT2i at the start of follow-up (N=44,286). Independent follow-ups were performed to identify the study outcomes – Cox regression to estimate adjusted hazard ratios (HRs) for the study outcomes: cardiovascular (CV) composite outcome (comprising non-fatal myocardial infarction [MI]/ischaemic stroke [IS] requiring hospitalisation and CV death), severe renal disease, and all-cause mortality.

**Results:** Mean follow-up was 2.3 years (SGLTi cohort) and 2.1 years (comparison cohort). Mean age was 60.4 years (SD ±10.2, SGLTi cohort) and 60.4 years (SD ± 10.0, comparison cohort). SGLT2i new users were associated with a reduced risk of the CV composite (HR 0.75, 95% CI: 0.61–0.93), severe renal disease (HR 0.55, 95% CI: 0.46– 0.67), and all-cause mortality (HR 0.56, 95% CI: 0.49–0.63), with risk reductions similar irrespective of baseline CKD. Reduced risks were seen for IS (HR 0.51, 95% CI: 0.36–0.74) but not MI (HR 0.98, 95% CI: 0.74–1.28). Results were consistent in sensitivity analyses.

**Conclusions:** In this population-based study, SGLT2is were associated with significant CV, renal and survival benefits among individuals with type 2 diabetes on metformin; the CV benefit was driven by a reduced risk of ischaemic stroke.

**What is already known on this topic?:**

- In randomized controlled trials (RCTs), sodium-glucose co-transporter-2 inhibitors (SGLT2is) and have shown good efficacy in reducing the risk of adverse cardiovascular (CV) and renal events in patients with type 2 diabetes.
- These benefits of SGLT2is have also been seen in observational studies, but have shown uncertainty around the evidence for benefits on myocardial infarction (MI).
- RCTs and observational studies differ in the characteristics of patients studied and in their adherence to treatment.

**What this study adds?:**

- In this matched retrospective cohort study among patients with type 2 diabetes using metformin, those who started an SGLT2i had significantly reduced risks of all-cause mortality (44% risk reduction), severe renal disease (50% risk reduction), a CV composite outcome (non-fatal MI/ischaemic stroke requiring hospitalisation/CV death; 25% risk reduction) and ischaemic stroke (49 risk reduction) compared with those who didn’t start a SGLT2i; however, the risk of non-fatal MI was not significantly different between groups.
- These findings indicate that the beneficial effects on CV disease seen in trials are driven by a reduced risk of ischaemic stroke.

**How this study might affect research, practice or policy:**

- These results confirm that the benefits of SGLT2i in patients with type 2 diabetes observed in clinical trials are applicable to real-world settings, thereby supporting an increasing role of SGLT2i in diabetes care.

## INTRODUCTION

Sodium-glucose co-transporter-2 inhibitors (SGLT2is) are a relatively new class of glucose-lowering medication that are increasingly being used to treat patients with type 2 diabetes. Although initially introduced as second-line treatment after metformin,^1^ these drugs have also shown efficacy in treating patients with renal and cardiovascular conditions. Large cardiovascular (CV) randomised controlled trials (RCTs) have shown that SGLT2is reduce the risk of hospitalisation due to heart failure or CV death in patients with type 2 diabetes by 23%.^2^ Accordingly, SGLT2is are now recommended in the European Society of Cardiology guidelines as a first-line treatment option for patients with type 2 diabetes who are either drug naïve or on metformin, and with atherosclerotic CV disease or at high CV risk.^3^ Furthermore, in April 2021 the United States Food and Drug Administration approved the use of SGLT2is to treat chronic kidney disease (CKD) in patients with or without type 2 diabetes.^4^

Some studies have explored the extent to which the effects of SGLT2is found in RCTs are seen in clinical practice.^5-10^ These have also generally found clear evidence that SGLT2is are associated with a reduced risk of mortality and adverse CV/renal outcomes when compared with other glucose-lowering drugs, although evidence relating to myocardial infarction is less certain.^5,9,10^ We aimed to provide further evidence on this topic by investigating whether the benefits of SGLT2is on adverse CV and renal outcomes, and mortality, in patients with type 2 diabetes, that are seen in clinical trials, can be reproduced in an observational study from the United Kingdom (UK) among patients taking metformin.

## RESEARCH DESIGN AND METHODS

### Study design and data source

We undertook a population-based comparative cohort study using data from the IQVIA Medical Research Data-UK (IMRD-UK) primary care database, formerly known as The Health Improvement Network. The database contains the anonymised electronic health records (EHRs) of approximately 6% of the UK population^11^ and the data held are representative of the UK in terms of age, sex and geographic distribution.^12^ Clinical and prescribing information are entered by primary care practitioners (PCPs) as part of routine patient care. Medical events (including diagnoses, hospital referrals, etc.) are recorded using Read codes,^13^ with a free text field that enables manual data entry for the addition of further details. Demographics, lifestyle factors and results of laboratory tests, including those for renal function (e.g. serum creatinine [SCr] values), are also recorded. Data from secondary care are entered into the patient’s primary care record retrospectively, and all prescriptions are automatically recorded upon issue. The study protocol was approved by an independent scientific research committee (reference SRC-19THIN059).

### Source population and cohort identification

Identification of the SGLT2 inhibitor (SGLT2i) and comparison cohorts is shown in **Supplementary Figure 1**. The source population included all individuals aged 20–89 years with a diagnosis of type 2 diabetes in the IMRD-UK and on treatment with metformin during the study inclusion period (1 January 2013 to 31 December 2018). Individuals in the source population were also required to have been registered with their PCP for at least 2 years and with no previous recorded prescription for a SGLT2i. From this source population we identified patients with a first SGLT2i prescription issued during the study period (N=19,300); the date of issue was the start date. For each member of the SGLT2i cohort, we used risk-set sampling to randomly select up to four individuals who had not been issued an SGLT2i prescription on the start date, matched on age, sex, frequency of PCP visits in the previous year, and previous use of other type of glucose-lowering medication, if any (sulfonylureas, dipeptidyl peptidase-4 inhibitors [DPP4is], glucagon-like peptide 1 receptor [GLP1] agonists, or insulin) – this cohort was the ‘comparison cohort’ (**Supplementary Figure 2**). Sampling was sequential without replacement, therefore the risk-set for a member of the SGLT2i cohort could include individuals who were, themselves, issued a first prescription for a SGLT2i at a later stage. When an individual was sampled (either as an SGLT2i initiator or as a matched non-initiator), they left the source population and were no longer eligible for sampling in future matched sets. There were 12,978 individuals in the SGLT2i cohort and 44,286 individuals in the comparison cohort. The start date for each member of the comparison cohort was the same as the state date for its matched SGLT2i initiator.

### Follow-up and study outcomes

Individuals in the two study cohorts were followed from the start date until the earliest of: the study outcome, death, age 90 years, or the end of the follow-up period (31 December 2019). Separate follow-ups were performed for each outcome of interest, excluding individuals with a record of that outcome before the start date. We evaluated three study outcomes: a composite CV outcome, severe renal disease, and all cause-mortality. The composite CV outcome included non-fatal myocardial infarction (MI), non-fatal ischaemic stroke IS (IS) requiring hospitalisation, and CV death. Severe renal disease was defined as an estimated glomerular filtration rate (eGFR) value <30 ml/min/1.73m^2^ (based on either eGFR values entered directly or eGFR values calculated from recorded SCr values using the CKD-EPI formula), or a Read code for end-stage renal disease (ESRD), renal replacement or dialysis.

### Other patient variables

Information on other patient variables was determined relative to the start date. We obtained data on patient demographics (age and sex), lifestyle variables, comorbidities, comedications and healthcare use. For lifestyle variables (body mass index, smoking, and alcohol intake) the most recent available record before the start date was obtained; individuals with no data in their EHR were allocated to a category ‘missing’ for that variable. Comorbidities were identified any time before the start date, and included CV disease and risk factors, cerebrovascular, respiratory, gastrointestinal, and neurological disease, diabetes-related complications (nephropathy and neuropathy), and major bleeding. We determined baseline renal function using the most recently recorded eGFR value in the year before the start date. We also determined baseline glycated haemoglobin (HbA1c), and albumin-to-creatinine ratio (ACR), based on the most recent value recorded in the year before the start date, as well as frailty using the electronic frailty index based on an algorithm developed and validated in the IMRD UK database.^14^ Co-medication use was determined from prescriptions issued before the start date, and included non-SGLT2i glucose-lowering medications (insulin, metformin, sulfonylureas, thiazolidinediones, meglitinides, GLP-1 agonists, and DPP-4is), and other relevant medications. Polypharmacy was defined as the number of different medications issued in the previous year. We also determined healthcare use based on the number of primary care visits, referrals, and hospitalisations in the year before the start date.

### Statistical analysis

Baseline characteristics of the study cohorts were described using frequency counts and percentages for categorical variables and means with standard deviation (SD) for continuous variables. Incidence rates of the study outcomes were calculated by dividing the number of observed cases by the respective total person-time, with 95% confidence intervals (CIs) estimated assuming a Poisson distribution. We fitted separate Cox proportional hazards regression models to estimate the hazard ratio (HR) for the outcome associated with SGLT2i use (vs. comparator), adjusted for all covariates as potential confounders, including CV and other comorbidities, co-medications, healthcare use, and lifestyle factors (see **Supplementary Table 1**). We used three different strategies of analysis. Firstly, we performed an intention-to-treat (ITT) analysis where we assumed that exposure status (SGLT2i or comparator) remained the same throughout follow-up – this approach was considered as the main analysis. Secondly, we performed an on-treatment (OT) analysis where we censored individuals if and when their initial exposure changed (i.e. discontinued SGLT2i for individuals in the SGLT2i cohort or initiated SGLT2i for individuals in the comparison cohort). Thirdly, we carried out an as-treated (AT) analysis where person-time was classified according to actual SGLT2i exposure during follow-up irrespective of the study cohort (that was based on initial exposure at the start date). Exposure to SGL2i in the AT analysis was categorised in relation to the time window after the end of the last SGLT2i consecutive prescription: *current use* (<30 days), *recent use* (between 30 and 90 days), *past use* (91– 365 days), and *non-use* (>365 days, or no previous prescription). We performed sensitivity analyses where we restricted analyses to individuals with eGFR values recorded in the year before the start of follow-up, and stratified results by CKD status at the start date. As differential mortality could influence the results, we performed further sensitivity analyses using Fine and Gray models to calculate adjusted sub-distribution HRs (SHRs) where death was considered a competing risk for the CV/renal outcomes.^15^ Analyses were undertaken using Stata version 12.1 (Statacorp).

## RESULTS

**Table 1** shows the characteristics of the two study cohorts at the start of follow-up. Mean age and sex distribution were similar in the SGLT2i cohort (59.6 years [SD±10.2], 39.1% female) and comparison cohort (60.4 years [SD±10.0], 39.1% females), as was the prevalence of CV diseases, diabetes-related complications, and frailty. However, notable differences were seen in the prevalence of CKD (9.0% in the SGLT2i cohort and 15.9% in the comparison cohort) and the proportion of patients with Hb1Ac over 8% at baseline (78.9% in the SGLT2i cohort and 34.9% in the comparison cohort). For each study outcome, the mean follow-up was 2.3 years for the SGLTi cohort and 2.1 years for the comparison cohort.

**Table 1.** Selected characteristics of the SGLT2i initiator and non-SGLT2i initiator (comparison) cohorts at the start of follow-up.

| **Variable** | **SGLT2i**<br>**N=12,978**<br>**n (%)** | **Non-SGLT2i**<br>**(comparison)**<br>**N=44,286**<br>**n (%)** |
| --- | --- | --- |
| **Age (years)** |  |  |
| 18–39 | 380 (2.9) | 975 (2.2) |
| 40–59 | 6179 (47.6) | 20,117 (45.4) |
| 60–79 | 6233 (48) | 22,477 (50.8) |
| ≥80 | 186 (1.4) | 717 (1.6) |
| Mean (SD) | 59.6 (10.2) | 60.4 (10.0) |
| **Sex** |  |  |
| Male | 7900 (60.9) | 27,557 (62.2) |
| Female | 5078 (39.1) | 16,729 (37.8) |
| **BMI (kg/m<sup>2</sup>)** |  |  |
| <20 | 26 (0.2) | 215 (0.5) |
| 20–24 | 541 (4.2) | 3856 (8.7) |
| 25–29 | 2862 (22.1) | 12,771 (28.8) |
| ≥30 | 9456 (72.9) | 26,978 (60.9) |
| Missing | 93 (0.7) | 466 (1.1) |
| **Smoking** |  |  |
| Non-smoker | 4929 (38) | 17,059 (38.5) |
| Current smoker | 1830 (14.1) | 7232 (16.3) |
| Ex-smoker | 6215 (47.9) | 19,976 (45.1) |
| Missing | 4 (0) | 19 (0) |
| **Alcohol use (units/week)** |  |  |
| None | 3264 (25.2) | 11,403 (25.7) |
| 1–9 | 6246 (48.1) | 20,485 (46.3) |
| 10–20 | 1645 (12.7) | 5792 (13.1) |
| 21–41 | 469 (3.6) | 1792 (4) |
| >41 | 236 (1.8) | 992 (2.2) |
| Missing | 1118 (8.6) | 3822 (8.6) |
| **Comorbidities** |  |  |
| Myocardial infarction | 804 (6.2) | 3098 (7) |
| IHD | 1899 (14.6) | 6976 (15.8) |
| CeVD | 744 (5.7) | 2974 (6.7) |
| Heart failure | 323 (2.5) | 1576 (3.6) |
| Hyperlipidaemia | 4120 (31.7) | 14,148 (31.9) |
| Hypertension | 7881 (60.7) | 27,142 (61.3) |
| Obesity | 4414 (34) | 12,732 (28.7) |
| CKD | 1170 (9.0) | 7059 (15.9) |
| Diabetic retinopathy | 4028 (31) | 14,090 (31.8) |
| Diabetic neuropathy | 272 (2.1) | 986 (2.2) |
| DKD | 291 (2.2) | 1062 (2.4) |
| **Frailty** |  |  |
| Fit | 3999 (30.8) | 13,971 (31.5) |
| Mild frailty | 6230 (48) | 19,714 (44.5) |
| Moderate frailty | 2266 (17.5) | 8258 (18.6) |
| Severe frailty | 483 (3.7) | 2343 (5.3) |
| **HbA1c** |  |  |
| >8% (>64 mmol/mol) | 10,240 (78.9) | 15,462 (34.9) |
| 7–8% (53–64 mmol/mol) | 2186 (16.8) | 12,743 (28.8) |
| <7% (<53 mmol/mol) | 410 (3.2) | 13,382 (30.2) |
| Missing | 142 (1.1) | 2699 (6.1) |
| **ACR (mg/g)** |  |  |
| Normal ($\leq 30$) | 5856 (45.1) | 18,593 (42) |
| Microalbuminuria ($>30$–$<300$) | 1715 (13.2) | 4932 (11.1) |
| Macroalbuminuria ($\geq 300$) | 211 (1.6) | 842 (1.9) |
| Missing | 5196 (40) | 19,919 (45) |
| **PCP visits<sup>†</sup>** |  |  |
| 0–12 | 2807 (21.6) | 11068 (25) |
| 13–24 | 6607 (50.9) | 21,499 (48.5) |
| 25–34 | 2242 (17.3) | 7198 (16.3) |
| $\geq 35$ | 1322 (10.2) | 4521 (10.2) |
| **eGFR (ml/min/1.73m<sup>2</sup>)** |  |  |
| <15 | 0 (0) | 3 (0) |
| 15–29 | 1 (0) | 148 (0.3) |
| 30–44 | 75 (0.6) | 1262 (2.8) |
| 45–59 | 550 (4.2) | 3648 (8.2) |
| 60–89 | 5711 (44) | 17701 (40) |
| $\geq 90$ | 6339 (48.8) | 18,216 (41.1) |
| Missing | 302 (2.3) | 3308 (7.5) |

<sup>†</sup>In the year before the start date (start of follow-up).
ACR, albumin-to-creatinine ratio; BMI, body mass index; CeVD, cerebrovascular disease; CKD, chronic kidney disease; DKD, diabetic kidney disease; eGFR, estimated glomerular filtration rate; IHD, ischaemic heart disease; PCP, primary care practitioner; SD, standard deviation; SGLT2i, sodium-glucose co-transporter-2 inhibitor

### Composite CV outcome

Results for the composite CV outcome of non-fatal MI/IS or CV death are shown in **Table 2**. The incidence rate of the composite CV outcome was 46.6 cases per 10,000 person-years (95% CI: 38.9–55.5) in the SGLT2i cohort, and 58.4 cases per 10,000 person-years (95% CI: 53.3–63.8) in the comparison cohort. Crude incidence rates were similar between the SGLT2i and comparison cohorts for non-fatal MI (30.5 vs. 30.0 cases per 10,000 person-years), and notably different for non-fatal IS (14.3 vs. 25.1 cases per 10,000 person-years). In the ITT analysis, after adjusting for confounders, the SGLT2i cohort had a 25% reduced risk of the composite CV outcome (HR 0.75, 95% CI: 0.61–0.93). A similar effect size was obtained with the Fine and Gray model (ITT analysis, SHR 0.77 (95% CI: 0.62–0.95) and in the AT analysis (HR 0.76, 95% CI: 0.61–0.95 for *current use*), while in the OT analysis, a greater reduced risk was seen (HR 0.54, 95% CI: 0.40–0.74). Compared with the comparison cohort, the SGLT2i cohort had a 46% reduced risk of IS (HR 0.51, 95% CI: 0.36–0.74) and no difference in the risk of MI (HR 0.98, 95% CI: 0.74–1.28). In stratified analyses, the CV benefits seen with use of SGLT2is were limited to individuals free of CKD at the start of follow-up: HRs for the CV composite were 0.70 (95% CI: 0.55–0.89) for no CKD, and 1.08 (95% CI: 0.67–1.73) for with CKD (**Supplementary Table 2**).

**Table 2.**
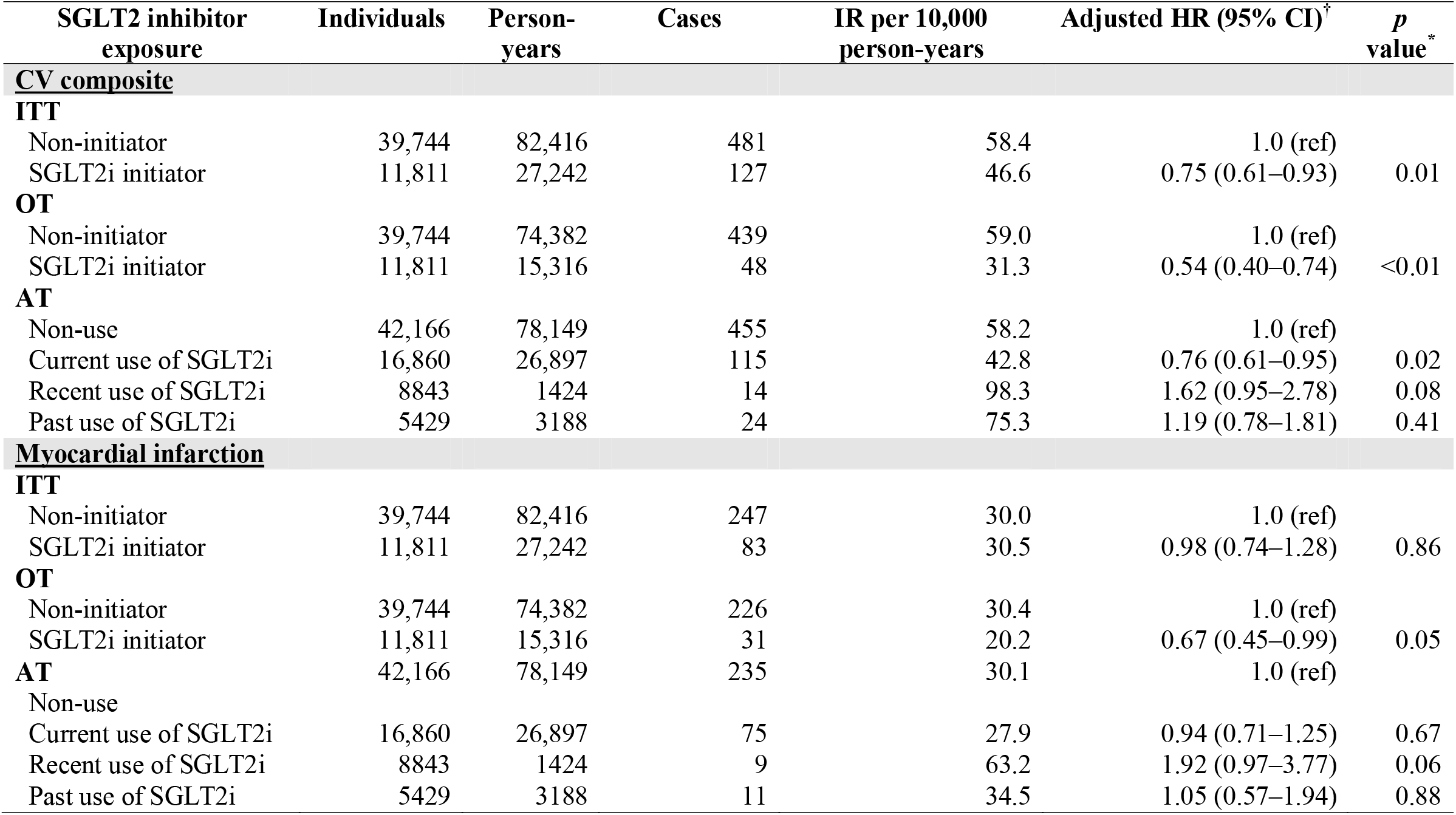
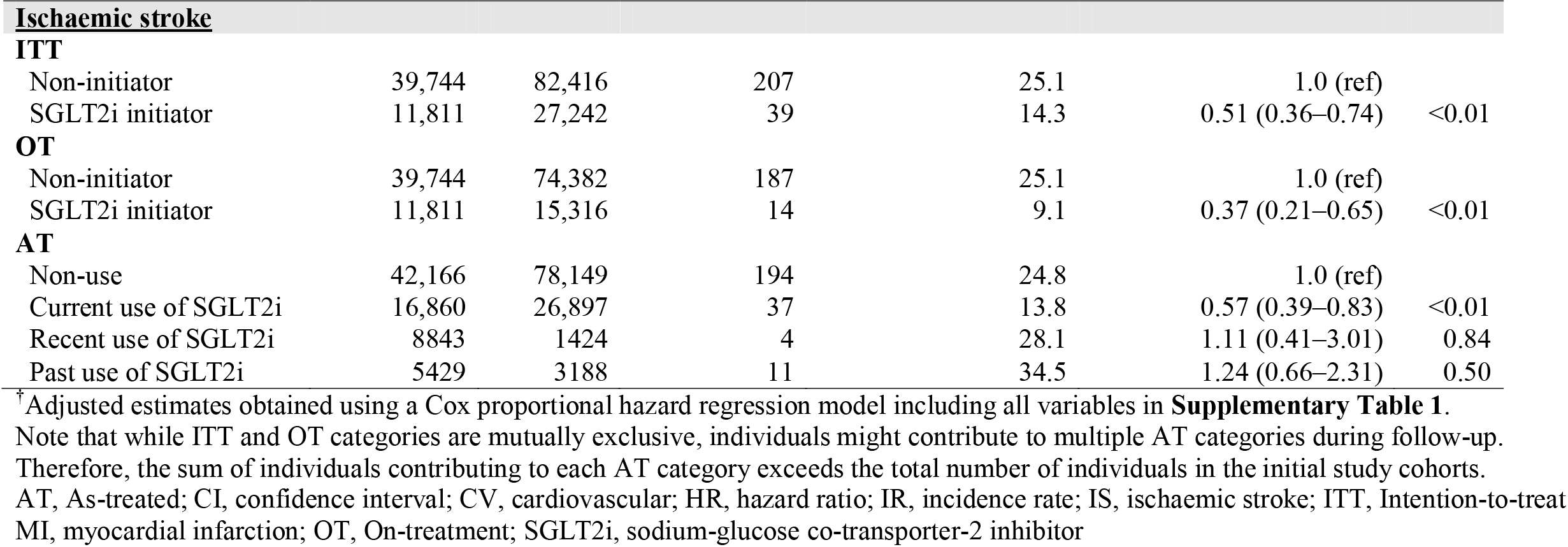
Fully adjusted HRs (95% CI) for MI, IS and the CV composite outcomes associated with SGLT2i initiation vs. non-SGLT2i initiation in patients with type 2 diabetes on metformin.

### Severe renal disease

Results for severe renal disease are shown in **Table 3**. The incidence rate of severe renal disease was 47.2 cases per 10,000 person-years (95% CI: 39.8–55.7) in the SGLT2i cohort, and 122.1 cases per 10,000 person-years (95% CI:115.0–129.5) in the comparison cohort. In the ITT analysis, we observed a 45% reduced risk of severe renal disease in the SGLT2i cohort, after adjusting for confounders (HR 0.55, 95% CI: 0.46– 0.67). The estimate from the Fine and Gray model was virtually identical (ITT analysis, SHR 0.57, 95% CI: 0.47–0.69). Risk estimates were lower in both the OT analysis (HR 0.34, 95% CI: 0.25–0.47), and the AT analysis (*current use*, HR 0.31, 95% CI: 0.24– 0.40). Incidence rates of severe renal disease varied by baseline CKD status, being 28.8 and 40.4 per 10,000 person-years in the SGLT2i and comparison cohorts free of CKD, compared with 232.3 and 594.6 per 10,000 person-years in the SGLT2i and comparison cohorts with CKD. Hazard ratios, however, were similar irrespective of baseline CKD status (**Supplementary Table 2**).

**Table 3.** Fully adjusted HRs (95% CI) for severe renal disease associated with SGLT2i initiation vs. non-SGLT2i initiation in patients with type 2 diabetes on metformin.

| SGLT2 inhibitor exposure | N | Person-years | Severe renal disease cases | IR per 10,000 person-years | Adjusted HR (95% CI)<sup>†</sup> | *p* value<sup>*</sup> |
| --- | --- | --- | --- | --- | --- | --- |
| **ITT** |  |  |  |  |  |  |
| Non-initiator | 43,852 | 90,437 | 1104 | 122.1 | 1.0 (ref) |  |
| SGLT2i initiator | 12,957 | 30,062 | 142 | 47.2 | 0.55 (0.46–0.67) | <0.01 |
| **OTT** |  |  |  |  |  |  |
| Non-initiator | 43,852 | 81,643 | 1076 | 131.8 | 1.0 (ref) |  |
| SGLT2i initiator | 12,957 | 16,933 | 43 | 25.4 | 0.34 (0.25–0.47) | <0.01 |
| **AT** |  |  |  |  |  |  |
| Non-use | 46,522 | 85,758 | 1131 | 131.9 | 1.0 (ref) |  |
| Current use of SGLT2i | 18,469 | 29,681 | 72 | 24.3 | 0.31 (0.24–0.40) | <0.01 |
| Recent use of SGLT2i | 9706 | 1565 | 10 | 63.9 | 0.70 (0.38–1.32) | 0.27 |
| Past use of SGLT2i | 5963 | 3495 | 33 | 94.4 | 0.82 (0.58–1.17) | 0.28 |

<sup>†</sup>Adjusted estimates obtained using a Cox proportional hazard regression model including all variables in **Supplementary Table**.
AT, As-treated; CI, confidence interval; HR, hazard ratio; IR, incidence rate; ITT, Intention-to-treat; OT, On-treatment; SGLT2i, sodium-glucose co-transporter-2 inhibitor
Note that while ITT and OT categories are mutually exclusive, individuals might contribute to multiple AT categories during follow-up. Therefore, the sum of individuals contributing to each AT category exceeds the total number of individuals in the initial study cohorts.

### All-cause mortality

Results for all-cause mortality are shown in **Table 4**. The mortality rate was 10.1 per 1000 person-years (95% CI: 9.0–11.3) in the SGLT2i cohort, and 19.4 per 1000 person-years (95% CI: 18.5–20.3) in the comparison cohort. After adjustment for confounders, we observed a 44% reduced mortality among the SGLT2i cohort (HR 0.56, 95% CI: 0.49–0.63). The reduced risk of death among the SGLT2i cohort was also seen in the OT analysis (HR 0.34, 95% CI: 0.27–0.42) and the AT analysis (*current use*, HR 0.35, 95% CI: 0.30–0.41). Mortality rates in the SGLT2i and comparison cohorts were 9.2 and 15.1 per 1000 person-years, respectively, among individuals free of CKD, compared with 21.1 and 41.3 per 1000 person-years, respectively, in individuals with CKD; however, HRs were similar in the stratified analysis by baseline CKD status (**Supplementary Table 2**).

**Table 4.**
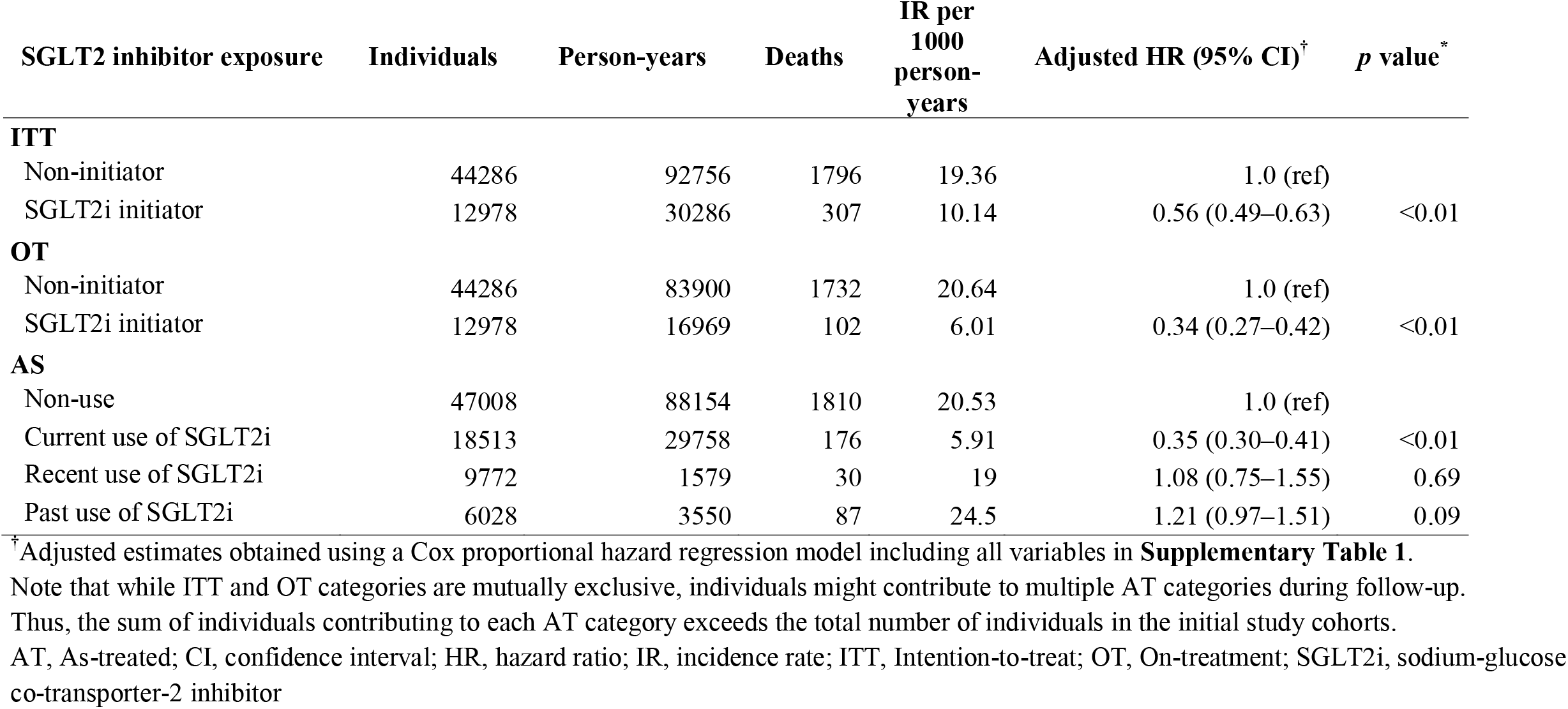
Fully adjusted HRs (95% CI) for all-cause mortality associated with SGLT2i initiation vs. non-SGLT2i initiation in patients with type 2 diabetes on metformin.

| SGLT2 inhibitor exposure | Individuals | Person-years | Deaths | IR per 1000 person-years | Adjusted HR (95% CI)<sup>†</sup> | *p* value<sup>*</sup> |
| --- | --- | --- | --- | --- | --- | --- |
| **ITT** |  |  |  |  |  |  |
| Non-initiator | 44286 | 92756 | 1796 | 19.36 | 1.0 (ref) |  |
| SGLT2i initiator | 12978 | 30286 | 307 | 10.14 | 0.56 (0.49–0.63) | <0.01 |
| **OT** |  |  |  |  |  |  |
| Non-initiator | 44286 | 83900 | 1732 | 20.64 | 1.0 (ref) |  |
| SGLT2i initiator | 12978 | 16969 | 102 | 6.01 | 0.34 (0.27–0.42) | <0.01 |
| **AS** |  |  |  |  |  |  |
| Non-use | 47008 | 88154 | 1810 | 20.53 | 1.0 (ref) |  |
| Current use of SGLT2i | 18513 | 29758 | 176 | 5.91 | 0.35 (0.30–0.41) | <0.01 |
| Recent use of SGLT2i | 9772 | 1579 | 30 | 19 | 1.08 (0.75–1.55) | 0.69 |
| Past use of SGLT2i | 6028 | 3550 | 87 | 24.5 | 1.21 (0.97–1.51) | 0.09 |

<sup>†</sup>Adjusted estimates obtained using a Cox proportional hazard regression model including all variables in **Supplementary Table 1**.
Note that while ITT and OT categories are mutually exclusive, individuals might contribute to multiple AT categories during follow-up.
Thus, the sum of individuals contributing to each AT category exceeds the total number of individuals in the initial study cohorts.
AT, As-treated; CI, confidence interval; HR, hazard ratio; IR, incidence rate; ITT, Intention-to-treat; OT, On-treatment; SGLT2i, sodium-glucose co-transporter-2 inhibitor

## DISCUSSION

In this population-based study of patients with type 2 diabetes using metformin, use of SGLT2is were associated with a 25% reduced risk of the composite CV outcome when compared with non-SGLT2i use, which was driven by a reduced risk of IS. Use of SGLT2is were also associated with a clear 44% reduction in the risk of all-cause mortality, and an almost 50% reduction in the risk of severe renal disease, irrespective of baseline CKD status.

Overall, our findings are consistent with the CV and renal benefits of SGLT2is seen both in RCTs that apply strict eligibility criteria, and previous observational studies. A recent meta-analysis based on six placebo-controlled trials RCTs found that SGLT2is were associated with a 10% reduction in the risk of major adverse CV events, and a 38% reduced risk of adverse kidney outcomes.^16^ A meta-analysis of cohort studies found SGLT2is to be associated with an 11% reduced risk of non-fatal IS vs. DPP4is, a non-significant 7% reduction in the risk of MI, and 26% reduced risk of all-cause mortality.^17^ In that study, the magnitude of the risk reductions were increased to 21% for stroke and 37% for all-cause mortality when pooling OT estimates rather than those from their ITT analysis, in line with the lower HRs seen in our OT analyses. A recent large international cohort study using data from claims, medical records and national registries reported a 51% reduced risk of renal events among individuals initiating SGLT2is vs. other glucose-lowering drugs.^5^ In another previous cohort study that compared SGLT2is with DPP4is using UK primary EHRs, Idris *et al*^10^ found the former to be associated with reduced risks of around 30% for all-cause mortality, 25% for CKD hospitalisation in patients with no history of CV/renal disease, and 51% for CKD hospitalisation in those with, or at high risk for, CVD; no reduction in risk seen for MI. In the US, Xie *et al*^9^ reported a 19% reduced risk of all-cause mortality among users of metformin taking SGLT2is vs. sulfonylureas as add-on therapy.

More than 460 million persons worldwide are currently living with diabetes, with type 2 accounting for around 90% of cases, and prevalence projected to increase.^18^ Furthermore, many individuals have prediabetes or undetected hyperglycaemia at the level of diabetes. Renal complications are strongly associated with an increased risk of stroke, MI, heart failure and mortality in persons with type 2 diabetes,^19-22^ and a key goal for diabetes care is to reduce organ injury and obtain a life-expectancy similar to persons without diabetes.^23^ It is therefore crucial to consider various treatment options based on the current evidence base, in addition to treatment costs.^24^ Among large RCTs of novel glucose-lowering agents on CV/ renal outcomes, different treatments have shown divergent effects.^25-29^ Although, no trial has performed a head-to-head comparison, several trials of DPP4is have failed to show any preventive renal effects.^25^ For GLP-1-analogues, a renal preventive effect of around 15%–20% has been shown, mainly due to effects on macroalbuminuria, but with no or little effect on renal function/ESRD.^26,27^ In contrast, SGLT2is have shown renal preventive effects, both in trials and real-world studies, and our present study suggests this effect is independent from baseline CKD status. As SGLT2is have also demonstrated a cardio-renal preventive effect in persons without diabetes, and in those with renal function <45 ml/min /1.73m^2^ where the glucose-lowering effect is reduced, these beneficial effects must act via other mechanisms.^30^ One likely mechanism is through lowering of the intraglomerular pressure, which is essential for preventing further renal progression.^30^ Moreover, renal complications are associated with increased activation of the renin-angiotensin-aldosterone system, altered metabolism and increased inflammation – all detrimental for cardiovascular complications.^31,32^

Although SGLT2is show a preventive effect on renal complications, it is important to note that renal function still progresses significantly in treated patients receiving this drug treatment.^5^ Furthermore, a small number of patients do not tolerate SGLT2is particularly well, and previous research has shown that, irrespective of CKD status, at least a quarter of patients with type 2 diabetes who are newly prescribed a SGLT2i discontinue their treatment within a year.^33^ Owing to the increased risk of ESRD, CVD and mortality in patients with impaired renal function, there is therefore a need for other complementary or alternative renal preventive treatments in type 2 diabetes care.Recently, finerenone, a drug that reduces mineralocorticoid receptor activation, has shown preventive effects on CKD progression and CVD in patients with type 2 diabetes and established CKD, and represents one such additional treatment option.^34,35^

Strengths of our study include the use large sample size from a data source representative of UK general population, hence our results have good generalisability. Also, as chronic disease such as diabetes, is largely managed in primary care, where all prescriptions are automatically recorded upon issue, medication use is likely to be well captured. We used different strategies of analysis – ITT, AT, and OT – as often used in clinical trials, as well as Fine and Gray models to account for the competing risk of death, and these showed our findings to be robust. One study limitation is the reliance on recorded eGFR values to classify individuals by baseline CKD status and to identify severe renal events during follow up. Although exposure to different glucose-lowering drugs could influence the frequency of renal function investigations, potentially biasing our results, virtually identical risk estimates were seen in the sensitivity analysis restricted to individuals whose renal function at baseline could be ascertained. One limitation is that renal function deterioration during follow-up could potentially affect SGLT2i use – either initiation or discontinuation of the drug for this very reason – and this would influence the results of the OT and AT analyses, where longitudinal change in drug use was considered. Also, it is not uncommon for people to discontinue life-extending medications in the last stages of life, and as we cannot exclude this as an explanation for the further reduced risk of mortality seen with current use of SGLT2is (as shown in the OT and AT analysis); caution is therefore needed when interpreting the time-dependent exposure mortality estimates.

In conclusion, our results indicate that among individuals with type 2 diabetes on metformin, use of SGLT2 is associated in significant CV, renal and survival benefits under normal conditions of use, confirming findings from RCTs on this topic. However, it is important to be aware of the level of unmet need that still exists in this patient population, as shown the high incidence of CV and renal outcomes in this present study, especially among those with concurrent CKD.

## Data Availability

All data produced in the present study are available upon reasonable request to the authors

## ACKNOWLEDGMENTS

We thank Susan Bromley of EpiMed Communications (Abingdon, UK), for medical writing assistance funded by Bayer AG in accordance with Good Publication Practice.

## DATA AVAILABILITY

Data are available from the corresponding author upon reasonable request.

## CONTRIBUTORSHIP

Study concept, DV, AG-P, LAGR; study design, AG-P, MES, LAGR, DV; data extraction and analysis; AG-P; interpretation of the data; all authors; review of manuscript drafts; all authors; final approval of the manuscript for publication, all authors.

## ETHICAL APPROVAL

The study protocol was approved by the Independent Scientific Research Committee for IMRD-UK (reference number reference reference SRC-19THIN059). Data collection for IMRD-UK was approved by the South East Multicentre Research Ethics Committee in 2003 and individual studies using IMRD-UK data do not require separate ethical approval if only anonymised data are used.

## REFERENCES

1. Davies MJ, D’Alessio DA, Fradkin J, et al. Management of hyperglycaemia in type 2 diabetes, 2018. A consensus report by the American Diabetes Association (ADA) and the European Association for the Study of Diabetes (EASD). Diabetologia 2018; 61(12): 2461–98.

2. Zelniker TA, Wiviott SD, Raz I, et al. SGLT2 inhibitors for primary and secondary prevention of cardiovascular and renal outcomes in type 2 diabetes: a systematic review and meta-analysis of cardiovascular outcome trials. Lancet 2019; 393(10166): 31–9.

3. Cosentino F, Grant PJ, Aboyans V, et al. 2019 ESC Guidelines on diabetes, pre-diabetes, and cardiovascular diseases developed in collaboration with the EASD. Eur Heart J 2020; 41(2): 255–323.

4. IQVIA. Is your Real World Data Credible? Part 2 in a series. How to identify the right set of data sources to address stakeholders’ broad evidence needs.

5. Heerspink HJL, Karasik A, Thuresson M, et al. Kidney outcomes associated with use of SGLT2 inhibitors in real-world clinical practice (CVD-REAL 3): a multinational observational cohort study. Lancet Diabetes Endocrinol 2020; 8(1): 27– 35.

6. Birkeland KI, Jørgensen ME, Carstensen B, et al. Cardiovascular mortality and morbidity in patients with type 2 diabetes following initiation of sodium-glucose co-transporter-2 inhibitors versus other glucose-lowering drugs (CVD-REAL Nordic): a multinational observational analysis. Lancet Diabetes Endocrinol 2017; 5(9): 709–17.

7. Kim YG, Han SJ, Kim DJ, Lee KW, Kim HJ. Association between sodium glucose co-transporter 2 inhibitors and a reduced risk of heart failure in patients with type 2 diabetes mellitus: a real-world nationwide population-based cohort study. Cardiovasc Diabetol 2018; 17(1): 91.

8. Kosiborod M, Lam CSP, Kohsaka S, et al. Cardiovascular Events Associated With SGLT-2 Inhibitors Versus Other Glucose-Lowering Drugs: The CVD-REAL 2 Study. J Am Coll Cardiol 2018; 71(23): 2628–39.

9. Xie Y, Bowe B, Gibson AK, McGill JB, Maddukuri G, Al-Aly Z. Comparative Effectiveness of Sodium-Glucose Cotransporter 2 Inhibitors vs Sulfonylureas in Patients With Type 2 Diabetes. JAMA Internal Medicine 2021; 181(8): 1043–53.

10. Idris I, Zhang R, Mamza JB, et al. Lower risk of hospitalization for heart failure, kidney disease and death with sodium-glucose co-transporter-2 inhibitors compared with dipeptidyl peptidase-4 inhibitors in type 2 diabetes regardless of prior cardiovascular or kidney disease: A retrospective cohort study in UK primary care. Diabetes, Obesity and Metabolism 2021; 23(10): 2207–14.

11. IQVIA. The Health Improvement Network (THIN).

12. Blak BT, Thompson M, Dattani H, Bourke A. Generalisability of The Health Improvement Network (THIN) database: demographics, chronic disease prevalence and mortality rates. Inform Prim Care 2011; 19(4): 251–5.

13. NHS Digital. Read codes.

14. Clegg A, Bates C, Young J, et al. Development and validation of an electronic frailty index using routine primary care electronic health record data. Age Ageing 2016; 45(3): 353–60.

15. Fine JP, Gray RJA. Proportional Hazards Model for the Subdistribution of a Competing Risk.. J Am Stat Assoc 1999; 94(446).

16. McGuire DK, Shih WJ, Cosentino F, et al. Association of SGLT2 Inhibitors With Cardiovascular and Kidney Outcomes in Patients With Type 2 Diabetes: A Meta-analysis. JAMA Cardiology 2021; 6(2): 148–58.

17. Mascolo A, Scavone C, Scisciola L, Chiodini P, Capuano A, Paolisso G. SGLT-2 inhibitors reduce the risk of cerebrovascular/cardiovascular outcomes and mortality: A systematic review and meta-analysis of retrospective cohort studies. Pharmacol Res 2021; 172: 105836.

18. International Diabetes Federation. IDF Diabetes Atlas. Ninth edition 2019.

19. González-Pérez A, Saez M, Vizcaya D, Lind M, Garcia Rodriguez L. Incidence and risk factors for mortality and end-stage renal disease in people with type 2 diabetes and diabetic kidney disease: a population-based cohort study in the UK. BMJ Open Diabetes Res Care 2021; 9(1).

20. Tancredi M, Rosengren A, Svensson AM, et al. Excess Mortality among Persons with Type 2 Diabetes. N Engl J Med 2015; 373(18): 1720–32.

21. Tancredi M, Rosengren A, Olsson M, et al. The relationship between three eGFR formulas and hospitalization for heart failure in 54 486 individuals with type 2 diabetes. Diabetes Metab Res Rev 2016; 32(7): 730–5.

22. Hedén Ståhl C, Lind M, Svensson AM, et al. Long-term excess risk of stroke in people with Type 2 diabetes in Sweden according to blood pressure level: a population-based case-control study. Diabet Med 2017; 34(4): 522–30.

23. American Diabetes Association. Introduction: Standards of Medical Care in Diabetes—2021. Diabetes Care 2021; 44Supplement 1): S1.

24. Buse JB, Wexler DJ, Tsapas A, et al. 2019 Update to: Management of Hyperglycemia in Type 2 Diabetes, 2018. A Consensus Report by the American Diabetes Association (ADA) and the European Association for the Study of Diabetes (EASD). Diabetes Care 2020; 43(2): 487–93.

25. Cornel JH, Bakris GL, Stevens SR, et al. Effect of Sitagliptin on Kidney Function and Respective Cardiovascular Outcomes in Type 2 Diabetes: Outcomes From TECOS. Diabetes Care 2016; 39(12): 2304–10.

26. Mann JFE, Ørsted DD, Brown-Frandsen K, et al. Liraglutide and Renal Outcomes in Type 2 Diabetes. New England Journal of Medicine 2017; 377(9): 839–48.

27. Gerstein HC, Colhoun HM, Dagenais GR, et al. Dulaglutide and renal outcomes in type 2 diabetes: an exploratory analysis of the REWIND randomised, placebo-controlled trial. Lancet 2019; 394(10193): 131–8.

28. Wanner C, Inzucchi SE, Lachin JM, et al. Empagliflozin and Progression of Kidney Disease in Type 2 Diabetes. New England Journal of Medicine 2016; 375(4): 323–34.

29. Mosenzon O, Wiviott SD, Cahn A, et al. Effects of dapagliflozin on development and progression of kidney disease in patients with type 2 diabetes: an analysis from the DECLARE-TIMI 58 randomised trial. Lancet Diabetes Endocrinol 2019; 7(8): 606–17.

30. Fioretto P, Zambon A, Rossato M, Busetto L, Vettor R. SGLT2 Inhibitors and the Diabetic Kidney. Diabetes Care 2016; 39 Suppl 2: S165–71.

31. Gansevoort RT, Correa-Rotter R, Hemmelgarn BR, et al. Chronic kidney disease and cardiovascular risk: epidemiology, mechanisms, and prevention. Lancet 2013; 382(9889): 339–52.

32. Go AS, Chertow GM, Fan D, McCulloch CE, Hsu CY. Chronic kidney disease and the risks of death, cardiovascular events, and hospitalization. N Engl J Med 2004; 351(13): 1296–305.

33. Ofori-Asenso R, Sahle BW, Chin KL, et al. Poor adherence and persistence to sodium glucose co-transporter 2 inhibitors in real-world settings: Evidence from a systematic review and meta-analysis. Diabetes Metab Res Rev 2021; 37(1): e3350.

34. Bakris GL, Agarwal R, Anker SD, et al. Effect of Finerenone on Chronic Kidney Disease Outcomes in Type 2 Diabetes. N Engl J Med 2020; 383(23): 2219–29.

35. Pitt B, Filippatos G, Agarwal R, et al. Cardiovascular Events with Finerenone in Kidney Disease and Type 2 Diabetes. N Engl J Med 2021; 385(24): 2252–63.

