## Supplementary files for "Cardiovascular and Renal Outcomes among Patients with Type 2 Diabetes using SGLT2 Inhibitors added to Metformin: A Population-Based Cohort Study from the United Kingdom"

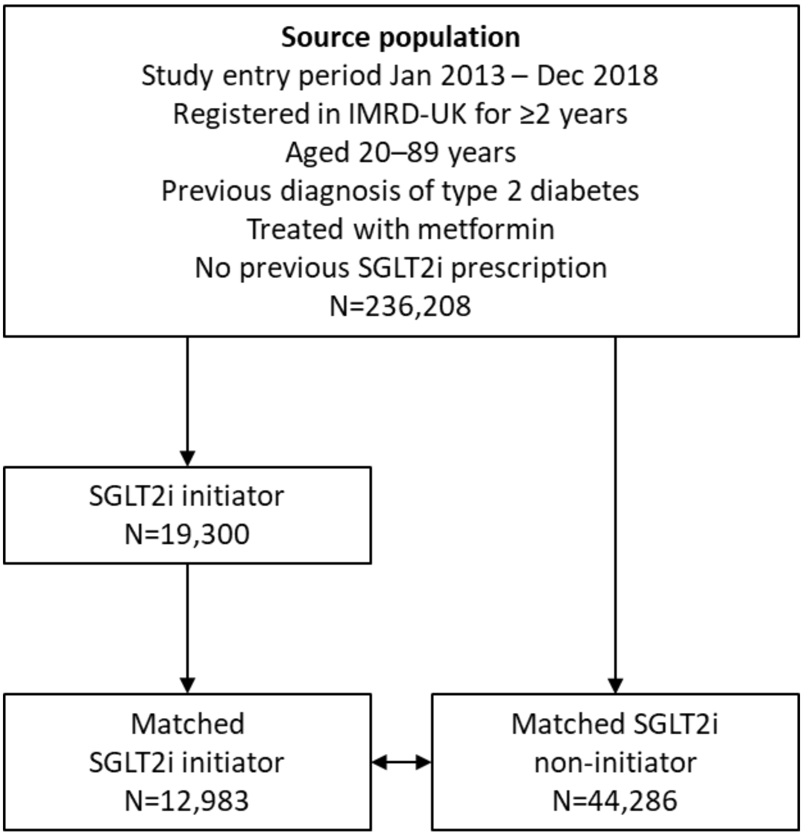


**Supplementary Figure 1.** Flowchart depicting identification of the SGLT2 inhibitor initiators cohort and the non-SGLT2i initiator cohort.
IMRD, IQVIA Medical Research Data UK; SGLT2, sodium-glucose co-transporter-2 inhibitor; UK, United Kingdom

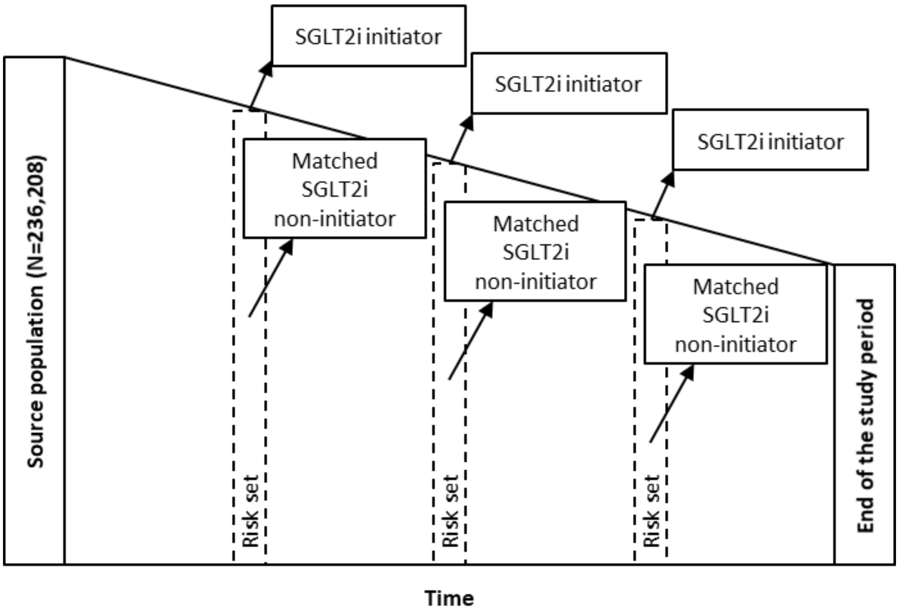


**Supplementary Figure 2.** Graphical description of matching procedure.
SGLT2, sodium-glucose co-transporter-2 inhibitor

**Supplementary Table 1**. Characteristics of the SGLT2i initiator and non-SGLT2i initiator (comparison) cohorts at the start of follow-up.

| **Variable** | **SGLT2i initiator N=12,978 n (%)** | **Non-SGLT2i initiator N=44,286 n (%)** |
|---|---|---|
| **Age (years)** | | |
| 18–39 | 380 (2.9) | 975 (2.2) |
| 40–59 | 6,179 (47.6) | 20,117 (45.4) |
| 60–79 | 6,233 (48.0) | 22,477 (50.8) |
| ≥80 | 186 (1.4) | 717 (1.6) |
| Mean (SD) | 59.6 (10.2) | 60.4 (10.0) |
| **Sex** | | |
| Male | 7,900 (60.9) | 27,557 (62.2) |
| Female | 5,078 (39.1) | 16,729 (37.8) |
| **BMI (kg/m$^2$)** | | |
| <20 | 26 (0.2) | 215 (0.5) |
| 20–24 | 541 (4.2) | 3,856 (8.7) |
| 25–29 | 2,862 (22.1) | 12,771 (28.8) |
| ≥30 | 9,456 (72.9) | 26,978 (60.9) |
| Missing | 93 (0.7) | 466 (1.1) |
| **Smoking** | | |
| Non-smoker | 4,929 (38.0) | 17,059 (38.5) |
| Current smoker | 1,830 (14.1) | 7,232 (16.3) |
| Ex-smoker | 6,215 (47.9) | 19,976 (45.1) |
| Missing | 4 (0.0) | 19 (0.0) |
| **Alcohol use (units/week)** | | |
| None | 3,264 (25.2) | 11,403 (25.7) |
| 1–9 | 6,246 (48.1) | 20,485 (46.3) |
| 10–20 | 1,645 (12.7) | 5,792 (13.1) |
| 21–41 | 469 (3.6) | 1,792 (4.0) |
| >41 | 236 (1.8) | 992 (2.2) |
| Missing | 1,118 (8.6) | 3,822 (8.6) |
| **Comorbidities** | | |
| Myocardial infarction | 804 (6.2) | 3,098 (7.0) |
| IHD | 1,899 (14.6) | 6,976 (15.8) |
| CeVD | 744 (5.7) | 2,974 (6.7) |
| Heart failure | 323 (2.5) | 1,576 (3.6) |
| History of ESRD | 5 (0.0) | 74 (0.2) |
| Hyperlipidaemia | 4,120 (31.7) | 14,148 (31.9) |
| Hypertension | 7,881 (60.7) | 27,142 (61.3) |
| Atrial fibrillation | 563 (4.3) | 2,225 (5.0) |

| Variable | SGLT2i initiator N=12,978 n (%) | Non-SGLT2i initiator N=44,286 n (%) |
|---|---|---|
| COPD | 771 (5.9) | 2,974 (6.7) |
| Cancer | 1,430 (11.0) | 5,617 (12.7) |
| Depression | 4,524 (34.9) | 14,720 (33.2) |
| Obesity | 4,414 (34.0) | 12,732 (28.7) |
| Acute pancreatitis | 199 (1.5) | 535 (1.2) |
| CKD | 1,170 (9.0) | 7,059 (15.9) |
| Chronic liver disease | 762 (5.9) | 2,399 (5.4) |
| Haemorrhagic stroke | 52 (0.4) | 257 (0.6) |
| Ischaemic stroke | 389 (3.0) | 1,727 (3.9) |
| TIA | 369 (2.8) | 1,345 (3.0) |
| Urogenital bleed | 1,331 (10.3) | 4,988 (11.3) |
| UGIB | 253 (1.9) | 1,016 (2.3) |
| LGIB | 1,102 (8.5) | 4,033 (9.1) |
| Liver disease | 1,217 (9.4) | 4,046 (9.1) |
| Pancreatic disease | 234 (1.8) | 713 (1.6) |
| Valvular disease | 208 (1.6) | 872 (2.0) |
| Tachycardia | 867 (6.7) | 3,332 (7.5) |
| PAD | 375 (2.9) | 1,552 (3.5) |
| Asthma | 2,761 (21.3) | 9,038 (20.4) |
| Rheumatoid arthritis | 422 (3.3) | 1,486 (3.4) |
| Osteoarthritis | 3,590 (27.7) | 12,425 (28.1) |
| Parkinson’s disease | 33 (0.3) | 149 (0.3) |
| Dementia psychosis | 67 (0.5) | 416 (0.9) |
| Alcohol abuse | 734 (5.7) | 2,955 (6.7) |
| IBD | 183 (1.4) | 654 (1.5) |
| Gastrointestinal disease | 9,266 (71.4) | 31,192 (70.4) |
| Diabetic retinopathy | 4,028 (31.0) | 14,090 (31.8) |
| Diabetic neuropathy | 272 (2.1) | 986 (2.2) |
| DKD | 291 (2.2) | 1,062 (2.4) |
| **$CHA_2DS_2VASc$ score** | | |
| 1 | 2,051 (15.8) | 7,129 (16.1) |
| 2 | 4,811 (37.1) | 15,202 (34.3) |
| 3 | 3,657 (28.2) | 12,495 (28.2) |
| 4 | 1,643 (12.7) | 6,057 (13.7) |
| 5 | 565 (4.4) | 2,324 (5.2) |
| 6 | 193 (1.5) | 801 (1.8) |
| 7 | 49 (0.4) | 237 (0.5) |
| ≥8 | 9 (0.1) | 41 (0.1) |

| **Variable** | **SGLT2i initiator N=12,978 n (%)** | **Non-SGLT2i initiator N=44,286 n (%)** |
|---|---|---|
| **Frailty** | | |
| Fit | 3,999 (30.8) | 13,971 (31.5) |
| Mild frailty | 6,230 (48.0) | 19,714 (44.5) |
| Moderate frailty | 2,266 (17.5) | 8,258 (18.6) |
| Severe frailty | 483 (3.7) | 2,343 (5.3) |
| **HbA1c** | | |
| >8% (>64mmol/mol) | 10,240 (78.9) | 15,462 (34.9) |
| 7–8% (53-64mmol/mol) | 2,186 (16.8) | 12,743 (28.8) |
| <7% (<53mmol/mol) | 410 (3.2) | 13,382 (30.2) |
| Missing | 142 (1.1) | 2,699 (6.1) |
| **ACR (mg/g)** | | |
| Normal (≤30) | 5,856 (45.1) | 18,593 (42.0) |
| Microalbuminuria (>30–<300) | 1,715 (13.2) | 4,932 (11.1) |
| Macroalbuminuria (≥300) | 211 (1.6) | 842 (1.9) |
| Missing | 5,196 (40.0) | 19,919 (45.0) |
| **PCP visits*** | | |
| 0–12 | 2,807 (21.6) | 11,068 (25.0) |
| 13–24 | 6,607 (50.9) | 21,499 (48.5) |
| 25–34 | 2,242 (17.3) | 7,198 (16.3) |
| ≥35 | 1,322 (10.2) | 4,521 (10.2) |
| **Referrals*** | | |
| 0–9 | 9,328 (71.9) | 31,895 (72.0) |
| 10–19 | 2,981 (23.0) | 9,917 (22.4) |
| ≥20 | 669 (5.2) | 2,474 (5.6) |
| **Hospitalisations*** | | |
| 0 | 10,963 (84.5) | 36,587 (82.6) |
| ≥1 | 2,015 (15.5) | 7,699 (17.4) |
| **eGFR (** ml/min/1.73m$^{2}$) | | |
| <15 | 0 (0.0) | 3 (0.0) |
| 15–29 | 1 (0.0) | 148 (0.3) |
| 30–44 | 75 (0.6) | 1,262 (2.8) |
| 45–59 | 550 (4.2) | 3,648 (8.2) |
| 60–89 | 5,711 (44.0) | 17,701 (40.0) |
| ≥90 | 6,339 (48.8) | 18,216 (41.1) |
| Missing | 302 (2.3) | 3,308 (7.5) |

| Variable | SGLT2i initiator N=12,978 n (%) | Non-SGLT2i initiator N=44,286 n (%) |
|---|---|---|
| **Polypharmacy** | | |
| 0–10 | 8,837 (68.1) | 33,495 (75.6) |
| 11–15 | 3,046 (23.5) | 8,025 (18.1) |
| ≥16 | 1,095 (8.4) | 2,766 (6.2) |
| **Previous use of other glucose-lowering drugs** | | |
| SU, GLP1, DDP4 and Insulin | 347 (2.7) | 891 (2.0) |
| GLP1, DDP4 and Insulin | 28 (0.2) | 33 (0.1) |
| SU, GLP1 and DDP4 | 563 (4.3) | 1,349 (3.0) |
| SU, DDP4 and insulin | 404 (3.1) | 1,214 (2.7) |
| SU, GLP1 and insulin | 452 (3.5) | 1,292 (2.9) |
| SU and Insulin | 468 (3.6) | 1,632 (3.7) |
| GLP1 and Insulin | 102 (0.8) | 179 (0.4) |
| GLP1 and DDP4 | 102 (0.8) | 150 (0.3) |
| SU and DDP4 | 3,013 (23.2) | 10,625 (24.0) |
| SU and GLP1 | 310 (2.4) | 722 (1.6) |
| DDP4 and insulin | 57 (0.4) | 77 (0.2) |
| SU only | 2,112 (16.3) | 7,992 (18.0) |
| Insulin only | 334 (2.6) | 1,003 (2.3) |
| GLP1 only | 90 (0.7) | 137 (0.3) |
| DDP4 only | 1,614 (12.4) | 5,160 (11.7) |
| None | 2,982 (23.0) | 11,830 (26.7) |
| **Warfarin** | | |
| Non-use | 12,547 (96.7) | 42,495 (96.0) |
| Current use | 403 (3.1) | 1,588 (3.6) |
| Past use | 28 (0.2) | 203 (0.5) |
| **Antiplatelets** | | |
| Non-use | 8,838 (68.1) | 29,633 (66.9) |
| Current use | 3,805 (29.3) | 13,384 (30.2) |
| Past use | 335 (2.6) | 1,269 (2.9) |
| **Parenteral anticoagulants** | | |
| Non-use | 12,927 (99.6) | 44,046 (99.5) |
| Current use | 9 (0.1) | 79 (0.2) |
| Past use | 42 (0.3) | 161 (0.4) |
| **Statins** | | |
| Non-use | 2,345 (18.1) | 8,790 (19.8) |
| Current use | 10,179 (78.4) | 33,521 (75.7) |
| Past use | 454 (3.5) | 1,975 (4.5) |
| **Antihypertensive drugs** | | |

| Variable | SGLT2i initiator N=12,978 n (%) | Non-SGLT2i initiator N=44,286 n (%) |
|---|---|---|
| Non-use | 3,193 (24.6) | 10,943 (24.7) |
| Current use | 9,555 (73.6) | 32,399 (73.2) |
| Past use | 230 (1.8) | 944 (2.1) |
| **Digoxin** | | |
| Non-use | 12,805 (98.7) | 43,692 (98.7) |
| Current use | 167 (1.3) | 555 (1.3) |
| Past use | 6 (0.0) | 39 (0.1) |
| **Antiarrhythmic drugs** | | |
| Non-use | 12,522 (96.5) | 42,591 (96.2) |
| Current use | 375 (2.9) | 1,400 (3.2) |
| Past use | 81 (0.6) | 295 (0.7) |
| **Aspirin** | | |
| Non-use | 9,274 (71.5) | 31,233 (70.5) |
| Current use | 3,330 (25.7) | 11,652 (26.3) |
| Past use | 374 (2.9) | 1,401 (3.2) |
| **Cox-2 inhibitors** | | |
| Non-use | 12,858 (99.1) | 43,959 (99.3) |
| Current use | 61 (0.5) | 177 (0.4) |
| Past use | 59 (0.5) | 150 (0.3) |
| **Paracetamol** | | |
| Non-use | 7,942 (61.2) | 27,471 (62.0) |
| Current use | 3,253 (25.1) | 10,806 (24.4) |
| Past use | 1,783 (13.7) | 6,009 (13.6) |
| **tNSAIDs** | | |
| Non-use | 10,387 (80.0) | 35,983 (81.3) |
| Current use | 1,079 (8.3) | 3,356 (7.6) |
| Past use | 1,512 (11.7) | 4,947 (11.2) |
| **PPIs** | | |
| Non-use | 7,660 (59.0) | 26,998 (61.0) |
| Current use | 4,297 (33.1) | 14,033 (31.7) |
| Past use | 1,021 (7.9) | 3,255 (7.3) |
| **H2 blockers** | | |
| Non-use | 12,481 (96.2) | 42,626 (96.3) |
| Current use | 377 (2.9) | 1,181 (2.7) |
| Past use | 120 (0.9) | 479 (1.1) |
| **Oral steroids** | | |
| Non-use | 11,967 (92.2) | 40,671 (91.8) |
| Current use | 387 (3.0) | 1,449 (3.3) |

| Variable | SGLT2i initiator N=12,978 n (%) | Non-SGLT2i initiator N=44,286 n (%) |
|---|---|---|
| Past use | 624 (4.8) | 2,166 (4.9) |
| **Inhaled steroids** | | |
| Non-use | 12,016 (92.6) | 40,961 (92.5) |
| Current use | 687 (5.3) | 2,476 (5.6) |
| Past use | 275 (2.1) | 849 (1.9) |
| **Injected steroids** | | |
| Non-use | 12,712 (98.0) | 43,492 (98.2) |
| Current use | 84 (0.6) | 171 (0.4) |
| Past use | 182 (1.4) | 623 (1.4) |
| **Anti-infective drugs** | | |
| Non-use | 6,436 (49.6) | 23,319 (52.7) |
| Current use | 2,330 (18.0) | 6,402 (14.5) |
| Past use | 4,212 (32.5) | 14,565 (32.9) |
| **Antidepressants** | | |
| Non-use | 9,051 (69.7) | 31,387 (70.9) |
| Current use | 3,248 (25.0) | 10,764 (24.3) |
| Past use | 679 (5.2) | 2,135 (4.8) |
| **Antipsychotics** | | |
| Non-use | 11,848 (91.3) | 40,296 (91.0) |
| Current use | 734 (5.7) | 2,527 (5.7) |
| Past use | 396 (3.1) | 1,463 (3.3) |
| **HRT** | | |
| Non-use | 12,819 (98.8) | 43,851 (99.0) |
| Current use | 104 (0.8) | 304 (0.7) |
| Past use | 55 (0.4) | 131 (0.3) |
| **Oral contraceptives** | | |
| Non-use | 12,752 (98.3) | 43,716 (98.7) |
| Current use | 173 (1.3) | 414 (0.9) |
| Past use | 53 (0.4) | 156 (0.4) |

*In the year before the start date (start of follow-up).
Note that medication use at start date was classified into current-use (less than 30 days from the end of the last available prescription), past-use (between 30 and 365 days from the end of the last available prescription), and non-use (more than 1 year since the end of the last prescription or no prior prescription).
ACR, albumin-to-creatinine ratio; BMI, body mass index; CeVD, cerebrovascular disease; CKD, chronic kidney disease; CLD, chronic liver disease; DDP4, dipeptidyl peptidase-4; DKD, diabetic kidney disease; eGFR, estimated glomerular filtration rate; ESRD, end-stage renal disease; GLP1, glucagon-like peptide-1; HRT, hormone replacement therapy; H2 blockers, histamine H2-receptor antagonists; IBD, inflammatory bowel disease; IHD, ischaemic heart disease; LGIB, lower gastrointestinal bleeding; PAD, peripheral artery disease; PCP, primary care practitioner; PPI, proton pump inhibitors; SD, standard deviation; SGLT2, sodium-glucose

co-transporter-2; SU, sulfonylurea; TIA, transient ischaemic attack; tNSAIDs, traditional nonsteroidal anti-inflammatory drug; UGIB, upper gastrointestinal disease bleeding

**Supplementary Table 2. Sensitivity analysis:** risk of study outcomes among individuals with available baseline eGFR, overall and stratified by baseline CKD status.

| | | | All individuals (with recorded eGFR)[*] | | | | | CKD (with recorded eGFR)[*] | | | | | No CKD (with recorded eGFR)[*] | | | | |
|---|---|---|---|---|---|---|---|---|---|---|---|---|---|---|---|---|---|
| | | **Exposure** | **N** | **Cases** | **IR per 1000 person-years** | **HR** | **95% CI** | **N** | **Cases** | **IR per 1000 person-years** | **HR** | **95% CI** | **N** | **Cases** | **IR per 1000 person-years** | **HR** | **95% CI** |
| **Mortality** | **ITT** | Non-initiator | 40,978 | 1680 | 19.53 | 1.0 | | 7048 | 607 | 41.26 | 1.0 | | 33,930 | 1073 | 15.05 | 1.0 | |
| | | SGLT2i initiator | 12,676 | 304 | 10.28 | 0.56 | (0.49–0.64) | 1170 | 58 | 21.06 | 0.54 | (0.40–0.72) | 11,506 | 246 | 9.17 | 0.56 | (0.48–0.65) |
| | **OT** | Non-initiator | 40,978 | 1620 | 20.84 | 1.0 | | 7048 | 597 | 42.32 | 1.0 | | 33,930 | 1023 | 16.07 | 1.0 | |
| | | SGLT2i initiator | 12,676 | 102 | 6.14 | 0.35 | (0.28–0.43) | 1170 | 20 | 15.07 | 0.42 | (0.27–0.67) | 11,506 | 82 | 5.37 | 0.33 | (0.26–0.42) |
| | **AT** | Non-use | 43,639 | 1696 | 20.72 | 1.0 | | 7446 | 613 | 41.60 | 1.0 | | 36,193 | 1083 | 16.13 | 1.0 | |
| | | Current use of SGLT2i | 17,832 | 173 | 6.02 | 0.36 | (0.30–0.42) | 1531 | 30 | 14.18 | 0.43 | (0.29–0.63) | 16,301 | 143 | 5.37 | 0.34 | (0.28–0.41) |
| | | Recent use of SGLT2i | 9432 | 30 | 19.69 | 1.11 | (0.77–1.60) | 943 | 2 | 12.66 | 0.31 | (0.08–1.27) | 8489 | 28 | 20.50 | 1.32 | (0.90–1.93) |
| | | Past use of SGLT2i | 5838 | 85 | 24.74 | 1.22 | (0.97–1.52) | 721 | 20 | 44.04 | 1.12 | (0.71–1.78) | 5117 | 65 | 21.80 | 1.21 | (0.94–1.57) |
| **Severe renal disease** | **ITT** | Non-initiator | 40,555 | 1047 | 12.50 | 1.0 | | 6625 | 760 | 59.46 | 1.0 | | 33,930 | 287 | 4.04 | 1.0 | |
| | | SGLT2i initiator | 12,655 | 138 | 4.70 | 0.57 | (0.47–0.69) | 1149 | 61 | 23.23 | 0.53 | (0.40–0.69) | 11,506 | 77 | 2.88 | 0.63 | (0.48–0.82) |
| | **OT** | Non-initiator | 40,555 | 1021 | 13.51 | 1 | | 6625 | 748 | 61.22 | 1.0 | | 33,930 | 273 | 4.31 | 1.0 | |
| | | SGLT2i initiator | 12,655 | 43 | 2.60 | 0.37 | (0.27–0.51) | 1149 | 21 | 16.16 | 0.44 | (0.28–0.70) | 11,506 | 22 | 1.44 | 0.30 | (0.19–0.48) |
| | **AT** | Non-use | 43,165 | 1074 | 13.50 | 1.0 | | 6994 | 773 | 60.53 | 1.0 | | 36,171 | 301 | 4.51 | 1.0 | |
| | | Current use of SGLT2i | 17,791 | 70 | 2.44 | 0.33 | (0.26–0.43) | 1494 | 30 | 14.54 | 0.38 | (0.26–0.55) | 16,297 | 40 | 1.50 | 0.31 | (0.22–0.44) |
| | | Recent use of SGLT2i | 9369 | 9 | 5.96 | 0.68 | (0.35–1.31) | 901 | 2 | 13.32 | 0.28 | (0.07–1.14) | 8468 | 7 | 5.14 | 1.16 | (0.55–2.48) |
| | | Past use of SGLT2i | 5776 | 32 | 9.46 | 0.83 | (0.58–1.19) | 682 | 16 | 37.83 | 0.73 | (0.44–1.21) | 5094 | 16 | 5.41 | 0.97 | (0.58–1.63) |
| **CV** | **ITT** | Non-initiator | 36,716 | 450 | 5.90 | 1.0 | | 5883 | 136 | 11.27 | 1.0 | | 30,833 | 314 | 4.89 | 1.0 | |
| | | SGLT2i initiator | 11,529 | 125 | 4.70 | 0.75 | (0.61–0.94) | 979 | 26 | 11.66 | 1.08 | (0.67–1.73) | 10,550 | 99 | 4.07 | 0.70 | (0.55–0.89) |
| | **OT** | Non-initiator | 36,716 | 412 | 5.99 | 1.0 | | 5883 | 131 | 11.34 | 1.0 | | 30,833 | 281 | 4.91 | 1.0 | |
| | | SGLT2i initiator | 11,529 | 48 | 3.20 | 0.55 | (0.40–0.76) | 979 | 9 | 8.13 | 0.80 | (0.38–1.67) | 10,550 | 39 | 2.81 | 0.50 | (0.35–0.72) |
| | **AT** | Non-use | 39,081 | 427 | 5.90 | 1.0 | | 6196 | 132 | 10.95 | 1.0 | | 32,885 | 295 | 4.89 | 1.0 | |

| | | | All individuals (with recorded eGFR)* | | | | | CKD (with recorded eGFR) * | | | | | No CKD (with recorded eGFR) * | | | | |
|---|---|---|---|---|---|---|---|---|---|---|---|---|---|---|---|---|---|
| | | **Exposure** | **N** | **Cases** | **IR per 1000 person-years** | **HR** | **95% CI** | **N** | **Cases** | **IR per 1000 person-years** | **HR** | **95% CI** | **N** | **Cases** | **IR per 1000 person-years** | **HR** | **95% CI** |
| | | Current use of SGLT2i | 16,228 | 112 | 4.31 | 0.77 | (0.61–0.96) | 1286 | 20 | 11.48 | 1.19 | (0.71–2.01) | 14,942 | 92 | 3.80 | 0.71 | (0.55–0.91) |
| | | Recent use of SGLT2i | 8520 | 13 | 9.48 | 1.56 | (0.89–2.73) | 774 | 2 | 15.69 | 1.34 | (0.32–5.60) | 7746 | 11 | 8.84 | 1.63 | (0.89–3.00) |
| | | Past use of SGLT2i | 5248 | 23 | 7.47 | 1.16 | (0.76–1.79) | 580 | 8 | 21.99 | 2.08 | (0.98–4.40) | 4668 | 15 | 5.53 | 0.95 | (0.56–1.61) |
| **MI** | **ITT** | Non-initiator | 36,716 | 230 | 3.02 | 1.0 | | 5883 | 64 | 5.31 | 1.0 | | 30,833 | 166 | 2.59 | 1.0 | |
| | | SGLT2i initiator | 11,529 | 82 | 3.09 | 0.99 | (0.75–1.30) | 979 | 18 | 8.08 | 1.38 | (0.75–2.54) | 10,550 | 64 | 2.63 | 0.90 | (0.66–1.24) |
| | **OT** | Non-initiator | 36,716 | 211 | 3.07 | 1.0 | | 5883 | 61 | 5.28 | 1.0 | | 30,833 | 150 | 2.62 | 1.0 | |
| | | SGLT2i initiator | 11,529 | 31 | 2.07 | 0.68 | (0.45–1.01) | 979 | 7 | 6.32 | 1.33 | (0.54–3.24) | 10,550 | 24 | 1.73 | 0.59 | (0.37–0.94) |
| | **AT** | Non-use | 39,081 | 220 | 3.04 | 1.0 | | 6196 | 62 | 5.14 | 1.0 | | 32,885 | 158 | 2.62 | 1.0 | |
| | | Current use of SGLT2i | 16,228 | 72 | 2.77 | 0.93 | (0.70–1.24) | 1286 | 16 | 9.18 | 1.82 | (0.96–3.43) | 14,942 | 56 | 2.31 | 0.81 | (0.58–1.12) |
| | | Recent use of SGLT2i | 8520 | 9 | 6.56 | 2.00 | (1.01–3.94) | 774 | 1 | 7.84 | 1.28 | (0.17–9.64) | 7746 | 8 | 6.43 | 2.17 | (1.05–4.46) |
| | | Past use of SGLT2i | 5248 | 11 | 3.57 | 1.06 | (0.57–1.97) | 580 | 3 | 8.24 | 1.39 | (0.42–4.64) | 4668 | 8 | 2.95 | 0.97 | (0.47–2.00) |
| **IS** | **ITT** | Non-initiator | 36,716 | 194 | 2.54 | 1.0 | | 5883 | 56 | 4.64 | 1.0 | | 30,833 | 138 | 2.15 | 1.0 | |
| | | SGLT2i initiator | 11,529 | 38 | 1.43 | 0.51 | (0.35–0.73) | 979 | 7 | 3.14 | 0.58 | (0.23–1.45) | 10,550 | 31 | 1.27 | 0.48 | (0.31–0.72) |
| | **OT** | Non-initiator | 36,716 | 176 | 2.56 | 1.0 | | 5883 | 54 | 4.67 | 1.0 | | 30,833 | 122 | 2.13 | 1.0 | |
| | | SGLT2i initiator | 11,529 | 14 | 0.93 | 0.38 | (0.21–0.67) | 979 | 1 | 0.90 | 0.13 | (0.02–1.13) | 10,550 | 13 | 0.94 | 0.39 | (0.21–0.71) |
| | **AT** | Non-use | 39081 | 182 | 2.51 | 1.0 | | 6196 | 54 | 4.48 | 1.0 | | 32,885 | 128 | 2.12 | 1.0 | |
| | | Current use of SGLT2i | 16,228 | 37 | 1.42 | 0.59 | (0.41–0.86) | 1286 | 3 | 1.72 | 0.35 | (0.10–1.24) | 14,942 | 34 | 1.40 | 0.62 | (0.42–0.93) |
| | | Recent use of SGLT2i | 8520 | 3 | 2.19 | 0.87 | (0.27–2.73) | 774 | 1 | 7.84 | 1.87 | (0.24–14.55) | 7746 | 2 | 1.61 | 0.69 | (0.17–2.81) |
| | | Past use of SGLT2i | 5248 | 10 | 3.25 | 1.16 | (0.61–2.23) | 580 | 5 | 13.74 | 3.61 | (1.32–9.86) | 4668 | 5 | 1.84 | 0.71 | (0.28–1.75) |

*Adjusted estimates obtained using a Cox proportional hazard regression model including all variables in **Supplementary table 1**.

AT, As-treated; CI, confidence interval; CKD, chronic kidney disease; HR, hazard ratio; IR, incidence rate; IS, ischaemic stroke; ITT, Intention-to-treat; OT, On-treatment; SGLT2i, sodium-glucose co-transporter-2 inhibitor
